# Lessons from a rapid systematic review of early SARS-CoV-2 serosurveys

**DOI:** 10.1101/2020.05.10.20097451

**Authors:** Niklas Bobrovitz, Rahul Krishan Arora, Tingting Yan, Hannah Rahim, Nathan Duarte, Emily Boucher, Jordan Van Wyk, Timothy Grant Evans

## Abstract

**Background:** As the world grapples with the COVID-19 pandemic, there is increasing global interest in the role of serological testing for population monitoring and to inform public policy. However, limitations in serological study designs and test standards raise concerns about the validity of seroprevalence estimates and their utility in decision-making. There is now a critical window of opportunity to learn from early SARS-CoV-2 serology studies. We aimed to synthesize the results of SARS-CoV-2 serosurveillance projects from around the world and provide recommendations to improve the coordination, strategy, and methodology of future serosurveillance efforts.

**Methods:** This was a rapid systematic review of cross-sectional and cohort studies reporting seroprevalence outcomes for SARS-CoV 2. We included completed, ongoing, and proposed serosurveys. The search included electronic databases (PubMed, MedRXIV, BioRXIV, and WHO ICTPR); five medical journals (NEJM, BMJ, JAMA, The Lancet, Annals of Internal Medicine); reports by governments, NGOs, and health systems; and media reports (Google News) from December 1, 2019 to May 1, 2020. We extracted data on study characteristics and critically appraised prevalence estimates using Joanna Briggs Institute criteria.

**Results:** Seventy records met inclusion criteria, describing 73 studies. Of these, 23 reported prevalence estimates: eight preprints, 14 news articles, and one government report. These studies had a total sample size of 35,784 and reported 42 prevalence estimates. Seroprevalence estimates ranged from 0.4% to 59.3%. No estimates were found to have a low risk of bias (43% high risk, 21% moderate risk, 36% unclear). Fifty records reported characteristics of ongoing or proposed serosurveys. Overall, twenty countries have completed, ongoing, or proposed serosurveys.

**Discussion:** Study design, quality, and prevalence estimates of early SARS-CoV2 serosurveys are heterogeneous, suggesting that the urgency to examine seroprevalence may have compromised methodological rigour. Based on the limitations of included studies, future serosurvey investigators and stakeholders should ensure that: i) serological tests used undergo high-quality independent evaluations that include cross-reactivity; ii) all reports of serosurvey results, including media, describe the test used, sample size, and sampling method; and iii) initiatives are coordinated to prevent test fatigue, minimize redundant efforts, and encourage better study methodology.

**Other:** PROSPERO: CRD42020183634. No third-party funding.

## Introduction

As the world grapples with the COVID-19 pandemic, interest in SARS-CoV-2 serology testing and immunity studies is increasing. Serology (also known as antibody) testing represents an opportunity to systematically monitor the spread of symptomatic and asymptomatic SARS-CoV-2 infection, identify disproportionately affected populations, and study protective immunity. Many policymakers, public health officials and employers are contemplating the role of serosurveillance in strategies to reopen society.^1^

Several countries and organizations - including the World Health Organization, with its Solidarity II program - have begun ramping up serological testing efforts.^2,3^ However, limitations in serological study designs and test standards raise concerns about the validity of seroprevalence estimates and their utility in decision-making. Shortfalls in the first wave of SARS-CoV-2 nucleic acid diagnostic testing (including poor test sensitivity and specificity, lack of distribution at scale, and inconsistent testing protocols), and the associated public outcry, serve as a cautionary tale for efforts to implement antibody-based screening.^4^

There is now a critical window of opportunity to learn from early SARS-CoV-2 serology studies. This paper presents results from a rapid up-to-date systematic review and synthesis of the results of SARS-CoV-2 serosurveillance projects from around the world and provides recommendations to improve the coordination, strategy, and methodology of future serosurveillance efforts.

## Methods

To conduct this “living” rapid review, we used abbreviated systematic review methods informed by Cochrane guidance.^5^

### Registration and Reporting

The protocol for this review was registered (PROSPERO: CRD42020183634). The full protocol can be found in Supplementary File 1. Reporting for this review conformed to the PRISMA checklist (Supplementary File 2).

### Search Strategy

A rapid systematic review was undertaken, searching for published and unpublished SARS-CoV-2 serosurveys from December 1, 2019 to May 1, 2020 in: electronic databases (PubMed, MedRXIV, BioRXIV, and WHO ICTPR); high-impact medical journals (NEJM, BMJ, JAMA, The Lancet, Annals of Internal Medicine); reports by governments, NGOs, and health systems; and media reports (Google News). The complete search strategy can be found in Supplementary File 1.

### Inclusion and Exclusion Criteria

See Table 1 for study inclusion criteria and Table 2 for study exclusion criteria.

**Table 1:** Inclusion criteria.

| **Table 1: Inclusion criteria** |  |
| --- | --- |
| *Characteristics* | *Criteria for inclusion* |
| Population | • Humans - any age |
| Condition | • Previous SARS-CoV-2 infection (a.k.a. novel coronavirus, COVID-19) |
| Types of evidence | • Proposed or ongoing sero-surveys – defined as the collection and testing of serum (or proxy such as oral fluid) specimens from a sample of a defined population over a specified period of time to estimate the prevalence of antibodies against SARS-CoV-2 as an indicator of immunity<sup>5</sup><br>• Cross-sectional and cohort study designs, with serum measurements at single time points or repeated at multiple time points<br>• Published or unpublished academic literature, grey literature, media reports, or press releases |
| Outcome measures | • Report or provide data to calculate seroprevalence estimates:<br>◦ Seropositive prevalence (proportion with detectable antibodies)<br>◦ Seronegative prevalence (proportion without detectable antibodies)<br>◦ Seroprotected prevalence (proportion above protective antibody threshold)<br>◦ Non-seroprotected prevalence (proportion with no detectable antibodies or below the protective antibody threshold)<br>◦ Count/proportion of a population screened/unscreened |
| Languages | • Any |

**Table 2:** Exclusion criteria.

| **Table 2: Exclusion criteria** |  |
| --- | --- |
| *Characteristics* | *Criteria for exclusion* |
| Population | • Non-human (e.g., *in silico*, animal, *in vitro*) |
| Condition | • Active SARS-CoV-2 infection (a.k.a. novel coronavirus, COVID-19)<br>• Presence of SARS-CoV-2 antigen |
| Types of evidence | • Focus on COVID-19, but unrelated to serosurveillance (e.g., viral properties, general information about COVID-19)<br>• Study designs other than cross-sectional or cohort design<br>◦ Case reports, case-control studies, evaluations of serological tests, reviews of serological studies<br>• Serological studies that only include patients with previously confirmed COVID-19 infection<br>• Serological study protocols without an implementation plan that includes a proposed region, sample size, and approximate start date |
| Outcome measures | • Only reports incidence or prevalence of serum SARS-CoV-2 antigen |

### Article Screening and Data Extraction

Pairs of reviewers pilot screened 50 articles and extracted 5 articles in duplicate. All subsequent screening and data extraction was completed by one reviewer and verified by a second. Data on study characteristics, participants, and prevalence estimates were extracted. Discrepancies were resolved by discussion.

### Critical Appraisal

The estimates were critically appraised using the Joanna Briggs Checklist for Prevalence Studies.^7^ Two authors applied the criteria independently and in duplicate. Discrepancies were resolved by discussion. Based on these criteria, an overall risk of bias assessment was provided (i.e., low, moderate, high, unclear). The criteria were used to assess the extent to which systematic bias may have been introduced, the nature of the potential bias, and the magnitude of the potential bias. See Supplementary File 1 for additional notes regarding how the checklist was adapted for use in this rapid review.

### Data Presentation

All data were presented on a publicly accessible online platform, which can be accessed at serotracker.com. We designed built-in filters that allow users to sort prevalence estimates by region (i.e., states, provinces) and population (i.e., age, sex, health care workers, long term care residents, people aged 65+, people with chronic diseases/multi-morbidity). To contextualize each prevalence estimate, the total number of confirmed cases per one million population for the country on the start date for the study was extracted from Worldometer’s COVID-19 tracker.^8^

## Results

A total of 1,845 titles/abstracts and 1,267 full-text articles were screened, of which 70 were included for analysis (Figure 1).

**Figure 1:**
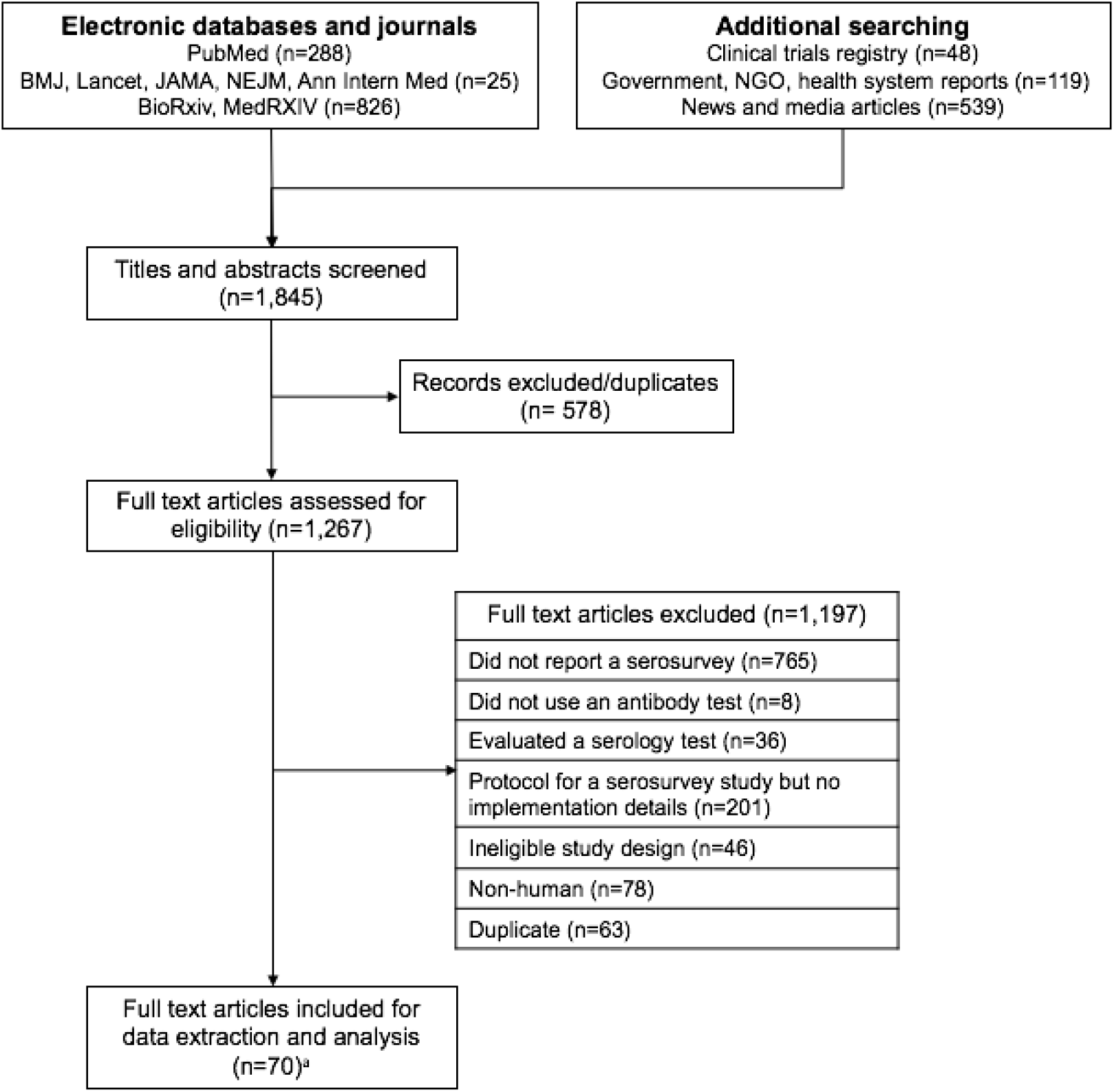
PRISMA Flow Diagram.

Seventy-three studies were found, reported in 70 articles. Twenty-three studies reported prevalence estimates (eight preprints, 14 news articles, and one government report). The vast majority of these studies (n=20/23, 87%) were in the United States and Europe. These studies had a total sample size of 35,784 and reported 42 prevalence estimates; 26 were final estimates and 16 were preliminary. Reported seroprevalence estimates ranged from 0.4% to 59.3% (Summary in Table 3; full results in Supplementary Table 5). Fifty additional studies were identified that intend to conduct 2.45 million tests and have not yet reported findings; most of these studies are also in the United States and Europe (Supplementary Table 6).

**Table 3:**
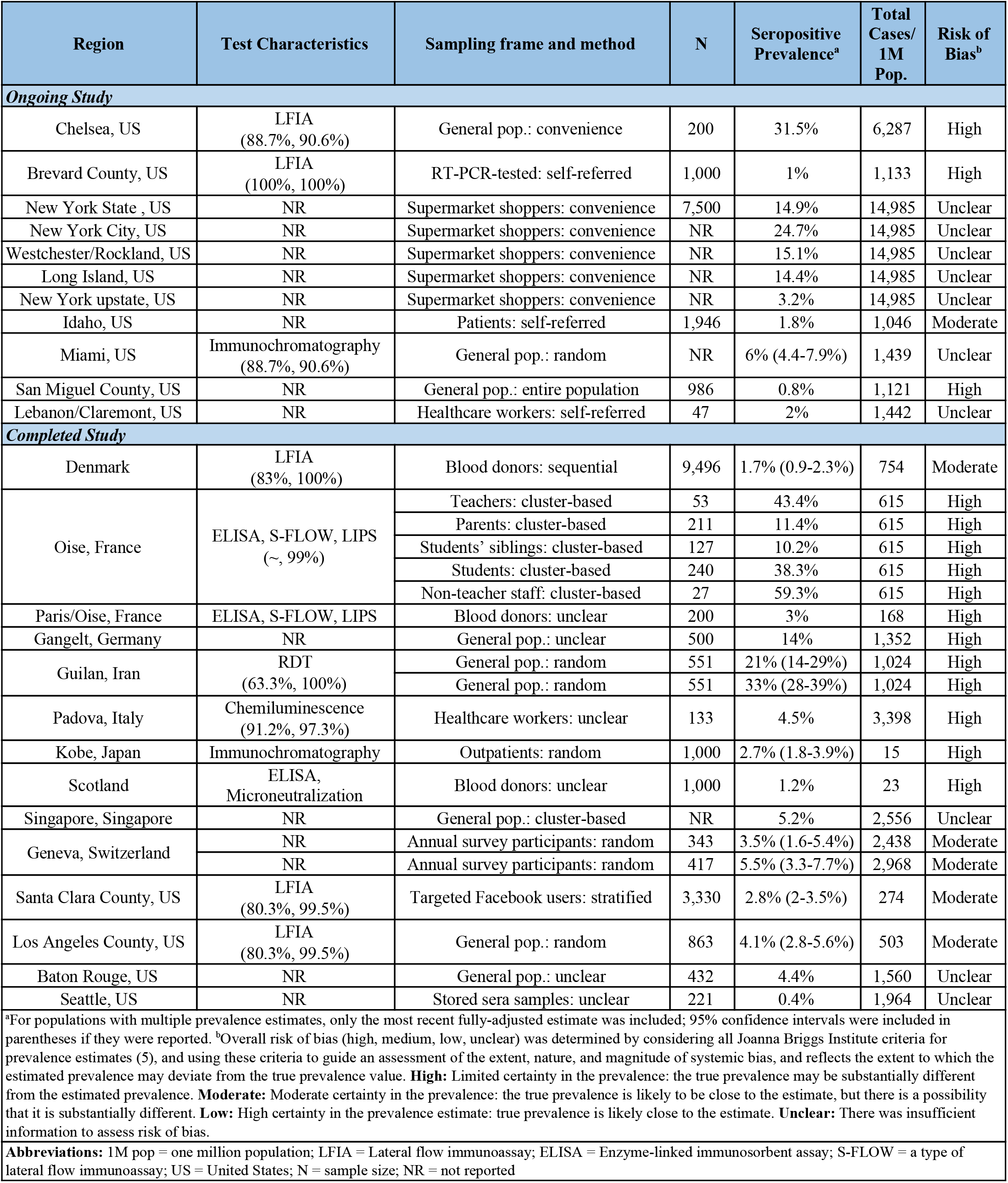
Reported prevalence estimates by region and population.

Overall, 14 countries have reported estimates from completed or ongoing studies, with six additional countries having proposed studies (Figure 2). Various stakeholders have taken the lead on these studies, including federal and regional governments, universities, health systems, businesses, and cooperative efforts (Table 4). Estimates used a range of test types, including ELISAs and lateral flow immunoassays (LFIAs), and examined a variety of antibodies including IgG alone, IgG and IgM, or IgG, IgM, and IgA.

**Figure 2:**
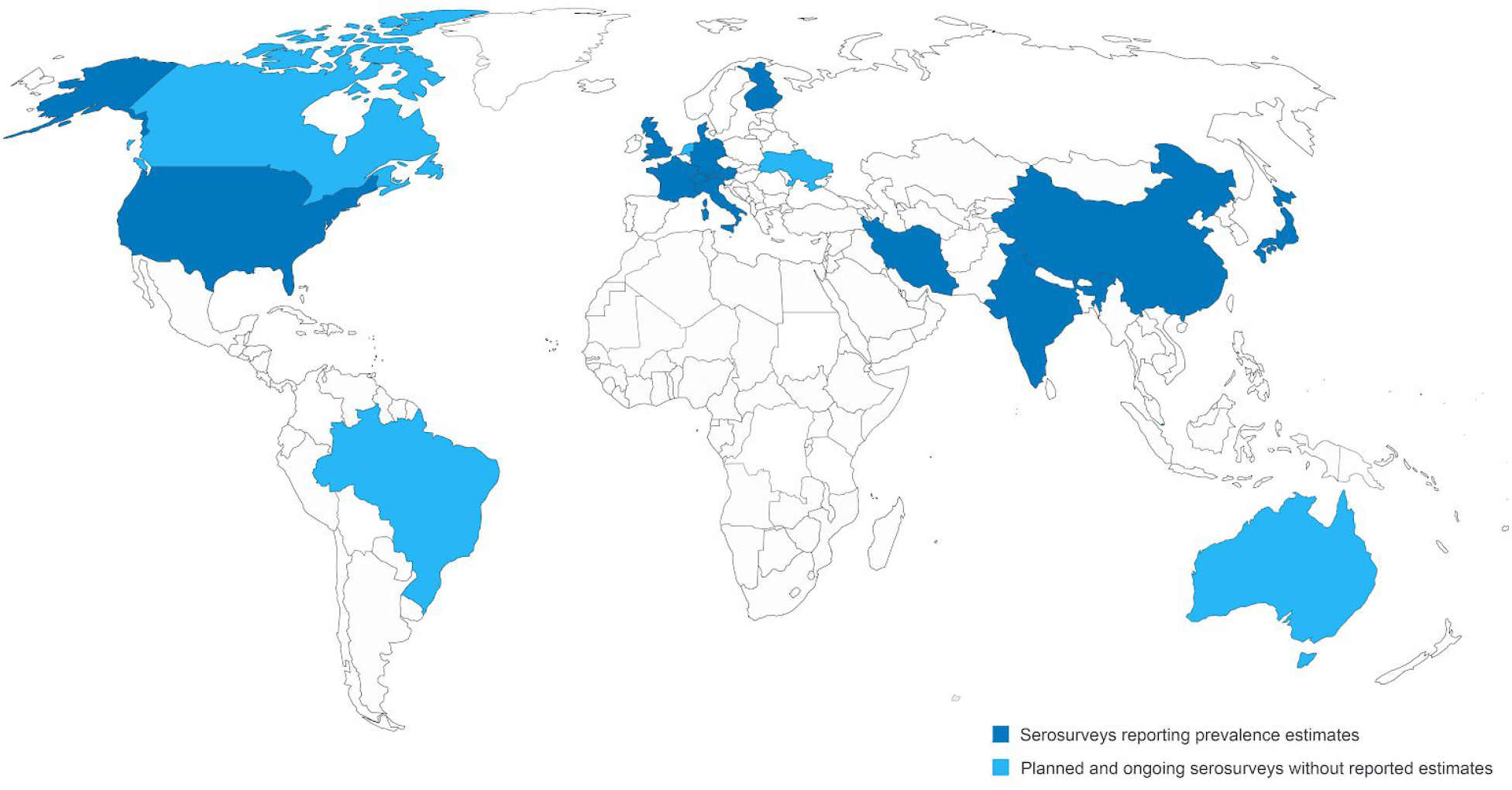
Map of countries with seroprevalence initiatives. Countries reporting data: Austria, China, Denmark, Finland, France, Germany, India, Iran, Italy, Japan, Singapore, Switzerland, United Kingdom, and the United States. Countries intending serosurveys but not yet reporting: Andorra, Australia, Brazil, Canada, Netherlands, and Ukraine.

**Table 4:** Organizations conducting serosurveys.

| Characteristic | n of studies (%) | Median sample size [min-max] |
| --- | --- | --- |
| ***For studies at all stages, including proposed (total intended n = 2,449,944)*** |  |  |
| Study status | 73 | 1,000 [27-1,000,000] |
| • Proposed | 21 (29%) | 25,000 [50-1,000,000] |
| • Ongoing | 38 (52%) | 1,000 [47-450,000] |
| • Completed | 14 (19%) | 432 [27-9,496] |
| ***For studies that have provided estimates (total recorded n = 35,784)*** |  |  |
| Organisation | 23 | 500 [27-9,496] |
| • One university or research institution | 5 (22%) | 282 [133-500] |
| • Multiple universities or research institutions | 5 (22%) | 396 [27-9,496] |
| • Health system | 3 (14%) | 432 [47-1,000] |
| • National institution | 1 (4%) | Not provided |
| • State institution | 4 (17%) | 2,473 [1,300-7,500] |
| • Private corporation | 1 (4%) | 986 [986-986] |
| • Collaboration across multiple types | 4 (17%) | 863 [200-3,330] |
| Risk of bias | 23 | 500 [27-9,496] |
| • High | 10 (43%) | 240 [27-1,000] |
| • Moderate | 5 (23%) | 1,405 [343-9,496] |
| • Low | 0 | n.a. |
| • Unclear | 9 (40%) | 866 [47-7,500] |

No estimates were found to have a low risk of bias (43% high risk, 21% moderate risk, 36% unclear). Study bias was predominantly attributed to inadequate sampling methods and antibody test performance (Supplementary Figure 3). Non-random sampling (e.g., self-referral) or a non-representative sampling frame (e.g., blood donors) characterized 61% of studies, and fewer than half of prevalence estimates were obtained from an appropriately sized sample (calculation in Appendix 1).^6^ Only two studies reported using tests with the United States FDA recommended minimum sensitivity and specificity (90% sensitivity, 95% specificity).^7^ The remainder used tests that failed to meet both thresholds (sensitivity 63.3% to 100%; specificity 90.63% to 100%) or failed to report test accuracy.

A full list of references for included studies is provided in Supplementary Table 7.

## Discussion

This rapid systematic review of SARS-CoV-2 serosurveys found twenty-three studies reporting data and fifty proposals for upcoming studies. Study design, quality, and results were heterogeneous. This snapshot of initial serosurveys suggests that the urgency to examine seroprevalence may have compromised methodological rigour and hence the validity of prevalence estimates. These early efforts to measure seroprevalence provide important insights on study design and coordination that will be critical to inform upcoming initiatives.

### Minimum standards of reporting

Serosurvey reports did not provide adequate information on their methods. Only 15 studies (65%) reported the test used, with only nine (39%) reporting test sensitivity and specificity. All reports of serosurveys, whether in published articles, preprints, grey literature or the news media, should meet minimum reporting standards.^9^ This includes the prevalence estimate itself, with confidence intervals and sample size; test name and characteristics; the dates and populations that these estimates apply to; and the sampling method.

Of course, the reporting standards for academic articles should be more rigorous. But in these unprecedented times, news articles are being used to rapidly report preliminary study results.^10^ It is therefore essential that these sources also provide the information needed to interpret their findings. If news agencies don’t hesitate to report on the findings of serological tests, then they shouldn’t hesitate to give their readers the complete picture.

### Serological test performance

While the advent of new immunoassays detecting antibodies to SARS-CoV-2 is welcome, the emergency conditions of the pandemic do not justify overlooking a thorough assessment of assay accuracy. As the recent experience in the UK reveals, rapid procurement of antibody tests only to find they are sub-standard represents an inefficient use of scarce public resources and risks undermining public confidence.^11^

Assessment of test characteristics relies on independent evaluation against a reference panel. Reference panels should include three key features: standardized positive and negative controls for the disease; a sufficient number of SARS-CoV-2 positive samples at different stages of infection, from day zero through 40+; as well as negative samples that have been exposed to other viruses (e.g., SARS-CoV-1, MERS and other viruses) to test for cross-reactivity.^12^

While independent evaluation of tests by a standards agency like the FDA is the norm, there is considerable variation in the way reference panels are prepared between agencies and hence standards are set. For example, the US FDA’s Emergency Use Authorization Pathway for serological tests requires the submission of test results on only 30 positive and 80 negative sera, and considers cross-reactivity against HIV but not other coronaviruses.^7^

Meanwhile, a variety of groups have conducted independent evaluations of SARS-CoV-2 antibody tests utilizing different volumes and types of sera in their reference panels.^13–15^ Investigators from UCSF included SARS-CoV-2 positive sera at multiple time points from infection and negative sera as part of the test evaluation for their study,^13^ while Danish study investigators included robust assessment of cross-reactivity with other coronaviruses and respiratory viruses.^14^ FINDDx is leading an ongoing independent evaluation program of 27 rapid diagnostic tests and seven ELISAs. Their evaluation should be commended for using large samples (100 positive and 100-300 negative sera) at multiple time points from symptom onset, but does not not appear to mandate cross-reactivity testing to ensure test specificity.^15^

Sensitivity and specificity thresholds for serology tests are a function of the prevalence of infection. As many have pointed out for initial seroprevalence results, 90% specificity may seem high, but can lead to as many false positives as true positives if prevalence is low. Insofar as evidence of infection correlates with decreased risk of re-infection - a variable of interest to guide decisions on re-opening society - there is a need to minimize false positives. As a result, the specificity threshold for SARS-CoV-2 antibody tests has moved towards 99%.

While the discussion of serology test characteristics has focused on specificity, sensitivity is also crucial. False negatives cannot be ignored, and are more likely in tests conducted soon after infection due to lower IgG antibody levels.^13^ There is a direct trade-off between sensitivity and specificity, and both are important to accurately assess population-level seroprevalence. To minimize false negatives, standards for sensitivity should also move towards 95%. Recent evaluations of commercial tests suggest that the market appears in a position to meet these higher specificity and sensitivity standards, which augurs well for serosurveys moving forward.

### Justify test choice and sampling method

Test selection can be complex and should be based on an overall testing strategy. Deciding between ELISAs, LFIAs, and other assay types requires considering the characteristics of the intended testing population. ELISAs require venous blood draws, making them easiest to deploy in hospitalized populations and long-term care facilities, and permit blood banking for future analysis.^16^ However, venipuncture may not be feasible in certain vulnerable populations, including people experiencing homelessness, in low-resource settings with poor access to care, or remote areas. LFIAs and paper blot assays are slightly less accurate, but are less invasive and less expensive alternatives that may better represent these groups.^16^

Few of the studies reviewed reported a rationale for their testing approach. Even of the eight included preprints, where one might expect the most thorough reporting, only three studies justified their choice of antibody test. When test choice was discussed, investigators and academics cited the lack of commercially available tests as a driving factor.

Less than half of studies so far have used an appropriate sample frame and random sampling method, with other studies using sampling designs that may oversample the young (recruiting through Facebook ads), healthy (blood donors), and high-SES groups (daytime drive-through testing).^17,18^ Adjusting estimates for population structure can provide a small degree of correction with large samples, but should not be substituted for strong sampling design. For example, the Santa Clara seroprevalence study weighted their sample by geography, sex, and race, but did not have a sufficiently large sample size to also weight by age, so seniors remained underrepresented in their weighted sample (4.5%) compared to the population (12.9%).^17^

In aggregate, there are few reports of seroprevalence in populations at high risk for SARS-CoV-2 infection, including lower socioeconomic status, older adults, people with comorbidities, and incarcerated populations. The first wave of studies was primarily focused on the general public, with a second wave of studies now emerging to study front line workers (e.g., health care providers, police officers).

COVID-19 has disproportionately affected marginalized populations; the UK and US are reporting higher in-hospital mortality rates for black and minority ethnic groups and seniors living in long term care facilities appear most at-risk everywhere.^19,20^ Moving forward, more appropriate sample frames tailored to the highly uneven and inequitable distribution of COVID-19 (as revealed by nucleic acid tests of acute infection) are needed to obtain more representative estimates of seroprevalence.

### The need for coordinated efforts

So far, serosurvey efforts have been scattered and partially overlapping. For example, the US National Institutes of Health is offering voluntary enrolment to citizens from all US states,^21^ overlapping with myriad state, municipality, university, and health system driven initiatives (Table 4) - each of which cover variable populations in terms of size and demographics.

Concurrently, private corporations such as Amazon and Barrick Gold are beginning to source hundreds of thousands of tests for their own employees, with no reported plans on how they will analyze and share seroprevalence data.^22^ Some test manufacturers are selling direct to consumer antibody test kits and private labs are offering pay-to-play testing, enabling people to self-refer for testing and potentially never disclose the result.

This lack of coordination could have two major consequences. Firstly, participants may develop test fatigue, wherein they are repeatedly offered antibody testing by multiple stakeholders and, after having received one result, refuse to participate in larger programs using higher quality tests that would yield more valuable and accurate results. Secondly, results obtained by a patchwork of stakeholders may never be reported and synthesized, meaning no one party will have a complete picture of the SARS-CoV-2 serology landscape.

Central registries of seroprevalence initiatives, and minimum regulatory standards on methodology and reporting, would coordinate efforts without stifling the autonomy of stakeholders eager to roll out antibody testing. These tools would make it possible to generate data that can be meaningfully compared and combined. They may also conserve resources by limiting duplication, allowing organisations to learn from each other, and integrating findings from nucleic acid and antibody testing to track active and past infections alike. Although central governance or partnerships can be slow to initiate, this disease may persist--and long-term problems require long-term solutions.

### Limitations

This review had some limitations. Firstly, it is possible that articles were missed by only searching one academic database of peer-review articles. That said, the supplemental search included five high-impact journals, two pre-print databases, and a trial registry. Secondly, we did not conduct article screening or extraction using two independent authors. However, we pilot-tested screening and extraction in duplicate to strengthen reliability. Furthermore, a second author verified screening decisions and extracted data.

### Conclusions

The world is entering the next phase of the SARS-CoV-2 pandemic - attempting a return to normalcy. The ability to accurately map seroprevalence patterns will be a key feature of this phase as scientists determine the relationship between antibody levels and immunity, and as decision-makers consider policies to ease restrictions on movement and reopen economies.

We should enter this phase armed with the lessons from early serosurveys: namely, that we need to raise the bar on seroprevalence testing initiatives and we need to do it together.

## Data Availability

All data is available at serotracker.com

https://serotracker.com/

## Acknowledgements

We thank Ewan May, Austin Atmaja, Simona Rocco, and Abel Joseph for their work to visualize findings from serosurveys and build serotracker.com.

## Ethics

No ethical approval was required for this work.

## Reporting

A PRISMA flow diagram and checklist have been included (Figure 1; Supplementary File 2).

